# Risk Factors for Adverse Outcomes with Outpatient Parenteral Antimicrobial Therapy

**DOI:** 10.1101/2023.04.21.23288837

**Authors:** Alexander J. Wang, Yaser Elnakieb, Esther Bae, Marguerite Monogue, James B. Cutrell, Christoph U. Lehmann, Christina Yen, Richard J. Medford

## Abstract

**Objectives:** Outpatient parenteral antimicrobial therapy (OPAT) use has increased significantly as it provides safe and reliable administration of long-term antimicrobials for severe infections. Benefits of OPAT include fewer antibiotic or line-related complications, increased patient satisfaction, shorter hospitalizations, and lower costs. Although OPAT programs carefully screen patients for eligibility and safety prior to enrollment, complications can occur. There is a paucity of studies identifying predictors of clinical outcomes in OPAT patients. Here, we seek to identify baseline predictors of OPAT outcomes utilizing machine learning methodologies.

**Methods:** We used electronic health record data from patients treated with OPAT between February 2019 and June 2022 at a large academic tertiary care hospital in Dallas, Texas. Three primary outcomes were examined: 1) clinical improvement at 30 days without evidence of reinfection; 2) patient actively being followed at 30 days; and 3) occurrence of any adverse event while on OPAT. Potential predictors were determined *a priori*, including demographic and clinical characteristics, OPAT setting, intravenous line type, and antimicrobials administered. Three classifiers were used to predict each outcome: logistic regression, random forest, and extreme gradient boosting (XGBoost). Model performance was measured using AUC, F1, and accuracy scores.

**Results:** We included 664 unique patients in the study, of whom 57% were male. At 30 days, clinical improvement was present in 78% of patients. Two-thirds of patients (67%) were actively followed at 30 days, and 30% experienced an adverse event while on OPAT. The XGBoost model performed best for predicting treatment success (average AUC = 0.873), with significant predictors including ID consultation and the use of vancomycin. The logistic regression model was best for predicting adverse outcomes (average AUC = 0.710). Risk factors for adverse outcomes included management in the home setting and the use of vancomycin, daptomycin, or piperacillin-tazobactam.

**Conclusion:** Outcomes of patients undergoing OPAT can be predicted with the use of easily-obtainable clinical and demographic factors. Patients requiring certain antimicrobial therapies, such as vancomycin or daptomycin, may derive less benefit from early hospital discharge and OPAT.

## INTRODUCTION

Outpatient parenteral antimicrobial therapy (OPAT) refers to the administration of intravenous (IV) antimicrobials to patients in ambulatory or community settings. OPAT is commonly prescribed to patients at discharge from the hospital and predominantly occurs at home. It may also be administered in skilled nursing facilities, inpatient rehabilitation centers, and dialysis centers. OPAT has been associated with shorter lengths of stay, lower hospitalization costs, and higher patient satisfaction.^1-5^

Despite the benefits, the lack of immediate, specialized supervision may heighten the risk of line malfunctions and incorrect dosing resulting in adverse events. With the use of OPAT expected to increase, it is important to identify risk factors associated with poor OPAT outcomes to mitigate preventable risks and better predict which patients may benefit from this therapy.^6,7^

## METHODS

### Study Setting and Design

The University of Texas Southwestern Medical Center (UTSW) is a large tertiary academic medical center and health system in Dallas, Texas, with an OPAT program that manages patients affiliated with its hospitals and ambulatory clinics. We conducted an observational cohort study using clinical and demographic data from patients enrolled in the OPAT program between February 10, 2019, and June 8, 2022. Data were collected and managed using REDCap (Research Electronic Data Capture), a secure, web-based platform designed to support data capture for research studies.^8^ The study was approved by the Institutional Review Board at UTSW.

Variables of interest were determined *a priori*, including demographics, diagnosis prompting OPAT enrollment, discharge setting, line type, OPAT team recommendations, antimicrobials used, treatment durations, laboratory results obtained for therapeutic drug monitoring, readmission details (if applicable), adverse drug reactions associated with OPAT, and 30- and 90-day outcome assessments.

We assessed three primary outcomes. The first outcome (M1) was clinical improvement of the patient without evidence of infection 30 days after completing OPAT. Patients who were lost to follow-up before completing therapy, experienced an infection relapse, were readmitted, or died due to infection or OPAT complications, were considered to have an unfavorable outcome. The second outcome (M2) was availability of data to evaluate the patient’s status 30 days after completion of therapy. Patients who were transferred to a different care system, were lost to follow-up after completing therapy, died for reasons unrelated to the infection, or were placed in hospice care, were considered “unable to be assessed.” All other patients were considered “assessable.” The final outcome (M3) was the occurrence of any adverse event while prescribed OPAT, identified as either an adverse drug reaction, central line complication, or readmission. This outcome was synthesized from existing data that would indicate one of the listed events.

### Data Processing

Our original data set contained information for each OPAT appointment. We combined appointment information for each patient and collapsed individual encounter data into unique patients. As different encounters may have recorded conflicting information for the same datum collected (e.g. a medication dose was changed), we generated rules to determine which datum was finally attributed to individual patients. For fields that contained the identical value for all encounters, we used that value. For fields with conflicting data, our goal was to propagate relevant values. For example, if a patient encounter had no socioeconomic issues recorded except for one occasion, we attributed the presence (rather than absence) of information to the patient.

For laboratory results, values were first converted to a binary normal or abnormal. There were multiple ways for a laboratory value to be considered normal. Most commonly, laboratory results within the range of accepted sex-specific normal values were considered normal. If a patient had the same laboratory test performed multiple times, the final data set was considered normal if all individual results were normal. If one result was abnormal, the data set was considered abnormal. Extreme (e.g. non-physiologic) laboratory values were assumed to be recordkeeping errors and marked as normal. Non-binary features, namely OPAT location/setting and ADR type, were merged through one-hot encoding, a conversion of categorical information into a format useable by machine learning algorithms. For each location and ADR type, we created a field that contained the count of occurrences.

Data on IV and oral antimicrobial administration were merged using similar methods. For patients, who had data on the intended duration of therapy but no actual duration recorded, the intended duration was substituted for the actual duration. Otherwise, intended durations were ignored. The merged patient data set contained each antimicrobial as a field, with each cell indicating the antimicrobial duration in days. We populated that field by summing the number of days a patient received each antimicrobial. Before adding an entry to the count, we ensured that entries were unique based on the combination of treatment start date and antimicrobial. Any fields with zero variance in values were removed.

### Modeling

We used three classifiers—logistic regression, random forest classification, and XGBoost—to model the three outcomes. Models M1 and M2 used all features. M3 included only features available prior to the 30-day outcome assessments. We included only patients with reported outcomes. For each classifier/outcome combination, we conducted a five-fold cross-validated grid search to find the optimal hyperparameters.

To reduce the number of irrelevant features, we applied Recursive Feature Elimination for each classifier/outcome combination using a step size of one and five-fold cross-validation. Various kernels were tested, with the best one selected for each combination. Because of the small size of the data set, different approaches to split data into training and testing groups caused significant variation in results. To account for this, we ran 100 executions for each classifier/outcome combination with unique train-test splitting and averaged the output. Each execution had a test size of 20% and 5-fold cross-validation. We determined optimal hyperparameters for each classifier/outcome combination using 5-fold cross-validated grid searches.

The number of features included in each model varied depending on outcome and RFECV kernel. Before feature selection, M1 and M2 included 156 features each, while M3 included 104. An XGBoost kernel, which cut M1 feature counts from 156 to 23, led to the best results for all classifiers. For M2, the optimal kernel varied by classifier, leading to feature counts between 30 and 37. The optimal kernel also varied for M3, leading to final feature counts between 40 and 50. The mean model coefficients for each feature were recorded, and the 95% confidence intervals were estimated using the 2.5^th^ percentile and 97.5^th^ percentile values across all executions.

For each classifier/outcome combination, one execution was chosen at random for displaying the receiver operating characteristic (ROC) and precision-recall (PR) curves. Because the test set sizes were small, we smoothed the curves using PCHIP monotonic interpolation between midpoints of vertical segments.

## RESULTS

The dataset included 3,114 individual encounters involving 664 unique patients. Of the 664 patients, 142 had a specified favorable or unfavorable 30-day outcome and were included in M1. An additional 71 patients did not have data from an assessment 30 days after therapy concluded. Thus, 213 patients were included in the model for M2. The model for M3 included all 664 patients, as its output was synthesized through adverse event data. Other demographic information is shown in **Table 1**.

**Table 1.** Patient characteristics.

| Variable | n (%) |
| --- | --- |
| Sex |  |
| Female | 289 (43.5) |
| Male | 375 (56.5) |
| Socioeconomic Status |  |
| Active Drug Use | 5 (0.75) |
| Historic Drug Use | 1 (0.15) |
| Unable to Afford OPAT | 19 (2.9) |
| Without Housing | 2 (0.3) |
| Other Issue(s) | 13 (2.0) |
| Infection Type |  |
| Bloodstream | 204 (30.7) |
| Bone and Joint | 209 (31.5) |
| Central Nervous System | 35 (5.3) |
| Diabetic Foot | 30 (4.5) |
| Endovascular | 74 (11.1) |
| Intra-abdominal | 67 (10.1) |
| Pulmonary | 25 (3.8) |
| Skin and Soft Tissue | 63 (9.5) |
| Surgical Site | 9 (1.4) |
| Urinary Tract | 61 (9.2) |
| Other Infection | 24 (3.6) |

The proportion of evaluable patients with favorable outcomes was 78% in M1. For M2, 67% of patients had available data on an assessment at 30 days, and the remaining were considered unable to be assessed. For the final outcome, M3, 30% of patients had one or more adverse events during therapy.

Overall model performance is summarized in **Table 2**. M1 had the highest area under the ROC curve (AUC), followed by M2 and M3. For M1, the XGBoost classifier provided the highest mean AUC (0.873), though the random forest model had higher F1 and accuracy scores. The logistic regression model performed best for M2 (mean AUC = 0.828) and M3 (mean AUC = 0.710).

**Table 2.** Overall model performance.

| 30-day outcomes (M1) |  |  |  |  |  |  |
| --- | --- | --- | --- | --- | --- | --- |
| Classifier | AUC Mean | AUC SD | F1 Mean | F1 SD | ACC Mean | ACC SD |
| Logistic Regression | 0.849 | 0.083 | 0.893 | 0.039 | 0.835 | 0.059 |
| Random Forest | 0.865 | 0.075 | 0.906 | 0.029 | 0.843 | 0.047 |
| XGBoost | 0.873 | 0.067 | 0.902 | 0.030 | 0.839 | 0.048 |
| Assessability at 30 days (M2) |  |  |  |  |  |  |
| Classifier | AUC Mean | AUC SD | F1 Mean | F1 SD | ACC Mean | ACC SD |
| Logistic Regression | 0.828 | 0.060 | 0.825 | 0.047 | 0.771 | 0.058 |
| Random Forest | 0.803 | 0.068 | 0.813 | 0.041 | 0.737 | 0.057 |
| XGBoost | 0.759 | 0.072 | 0.793 | 0.047 | 0.718 | 0.059 |
| Adverse events (M3) |  |  |  |  |  |  |
| Classifier | AUC Mean | AUC SD | F1 Mean | F1 SD | ACC Mean | ACC SD |
| Logistic Regression | 0.710 | 0.044 | 0.390 | 0.064 | 0.733 | 0.026 |
| Random Forest | 0.677 | 0.045 | 0.363 | 0.072 | 0.727 | 0.028 |
| XGBoost | 0.681 | 0.045 | 0.388 | 0.071 | 0.715 | 0.028 |

AUC SD = AUC standard deviation over 100 executions; ACC = model accuracy

The most important XGBoost features for M1 are shown in **Figure 1**. The use of an infectious disease team consultation during readmission, recurrent infection, and the antibiotics vancomycin and doxycline were the features that contributed most strongly to prediction of 30-day outcomes.

**Figure 1.**
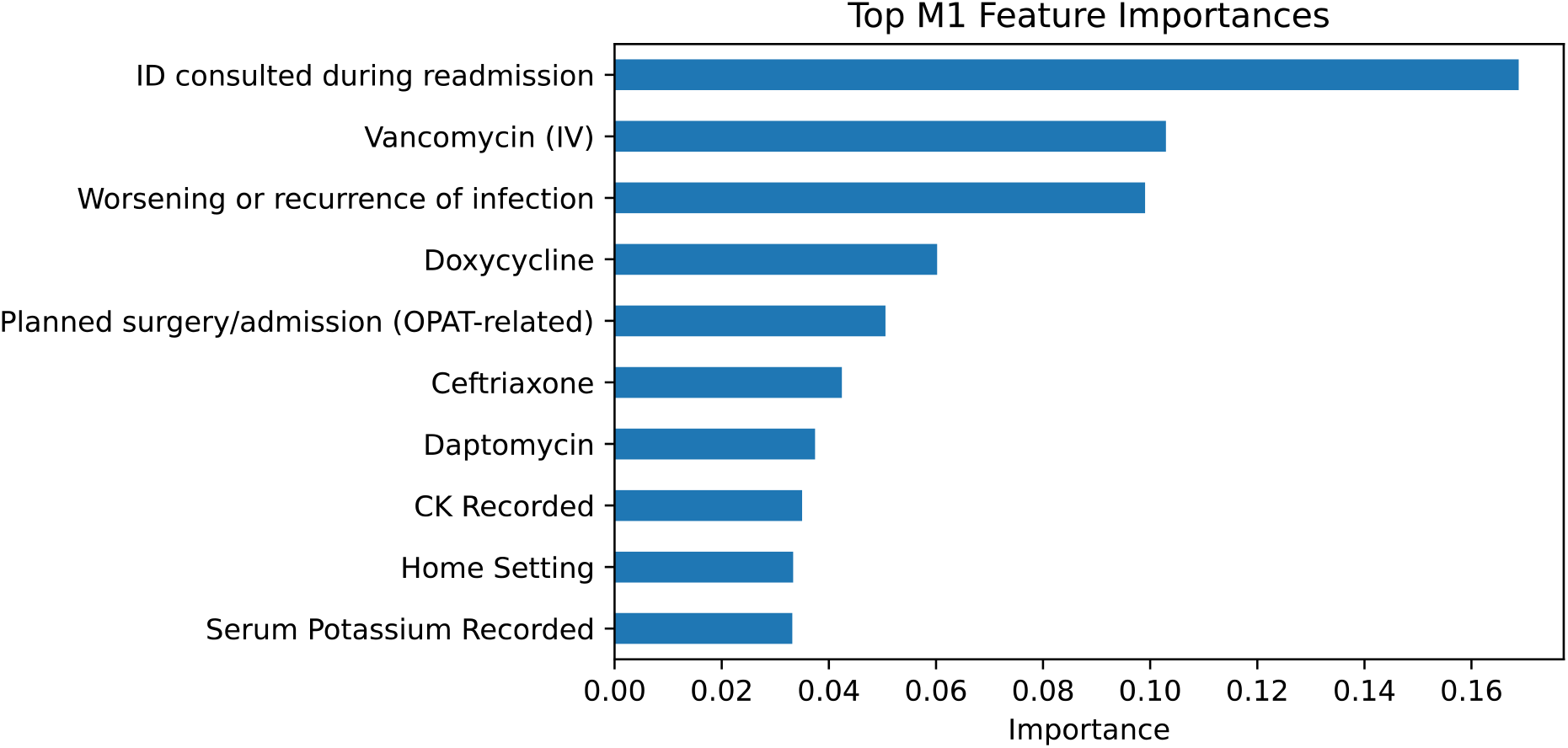
Features associated with outcomes at 30 days in the XGBoost model (M1).

The variables associated with a higher or lower odds of assessability at 30-days and adverse events are shown in **Tables 3 and 4**, respectively. Patients readmitted to the hospital, with recurrent infection, and/or treated at home were more likely to be assessable at 30 days. Those treated at home or administered vancomycin, daptomycin, or piperacillin-tazobactam were more likely to have an adverse event on therapy.

**Table 3.** Variables associated with assessability at 30 days (M2).

| Feature | Coefficient | Odds Ratio | 95% CI |
| --- | --- | --- | --- |
| Readmitted while on OPAT | 0.091 | 1.095 | 1.069, 1.122 |
| Readmission unrelated to OPAT | 0.084 | 1.088 | 1.065, 1.112 |
| Microbial culture/susceptibility pending | 0.073 | 1.075 | 1.055, 1.096 |
| Worsening or recurrence of infection | 0.064 | 1.066 | 1.048, 1.093 |
| Home setting | 0.058 | 1.060 | 1.028, 1.083 |
| ... | ... | ... | ... |
| Insufficient financial coverage | -0.060 | 0.942 | 0.911, 0.966 |
| Ampicillin | -0.068 | 0.934 | 0.907, 0.971 |
| Interventions prior to OPAT start | -0.078 | 0.925 | 0.903, 0.947 |
| Nursing home setting | -0.079 | 0.924 | 0.912, 0.940 |
| Follow-up imaging is required | -0.127 | 0.880 | 0.860, 0.907 |

CI: confidence interval. The 5 features with the highest coefficients and the 5 features with the lowest coefficients are shown.

**Table 4.** Variables associated with adverse events (M3).

| Feature | Coefficient | Odds Ratio | 95% CI |
| --- | --- | --- | --- |
| Home setting | 0.160 | 1.173 | 1.135, 1.208 |
| Piperacillin-tazobactam | 0.107 | 1.113 | 1.071, 1.153 |
| Vancomycin | 0.102 | 1.107 | 1.069, 1.144 |
| Daptomycin | 0.093 | 1.098 | 1.064, 1.141 |
| Bloodstream infection | 0.089 | 1.093 | 1.054, 1.314 |
| ... | ... | ... | ... |
| Skin and soft tissue infection | -0.071 | 0.931 | 0.905, 0.956 |
| Oral absorption/tolerance | -0.073 | 0.929 | 0.918, 0.945 |
| Endovascular infection | -0.081 | 0.922 | 0.895, 0.944 |
| Follow-up imaging is required | -0.103 | 0.902 | 0.873, 0.922 |
| No follow-up items | -0.129 | 0.879 | 0.853, 0.906 |

CI: confidence interval. The 5 features with the highest coefficients and the 5 features with the lowest coefficients are shown.

Randomly selected ROC and PR curves for each model and classifier are displayed in **Figures 2 and 3**.

**Figure 2.**
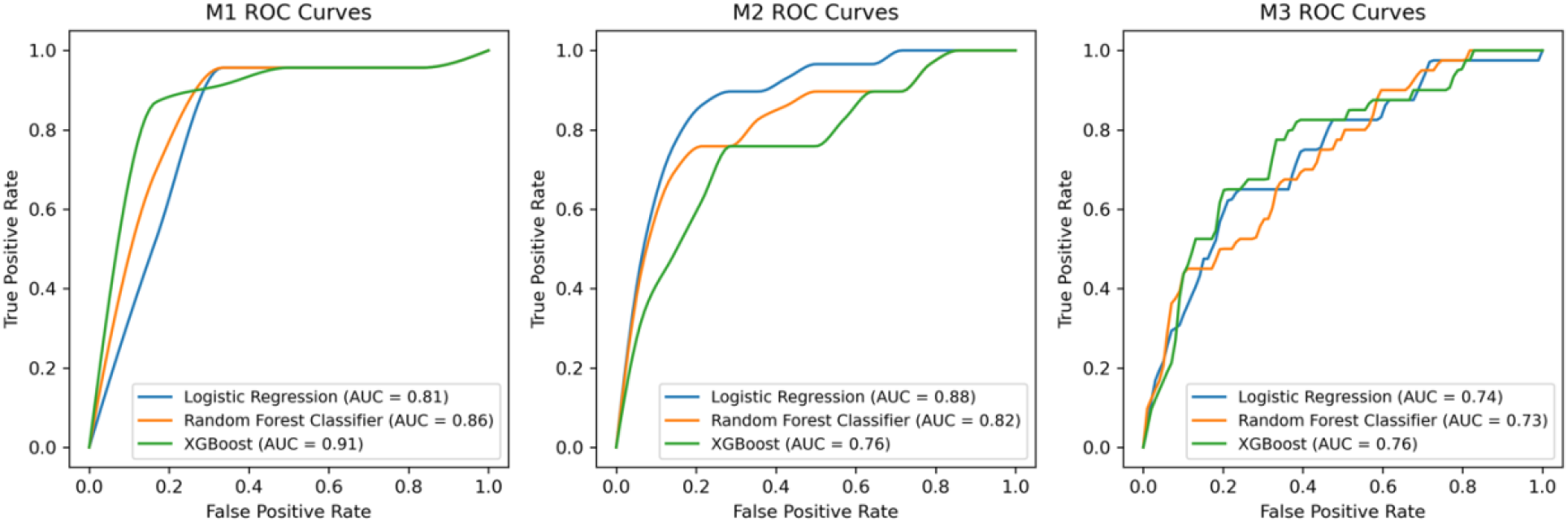
Receiver operating characteristics (ROC) curves, by outcome and classifier.

**Figure 3.**
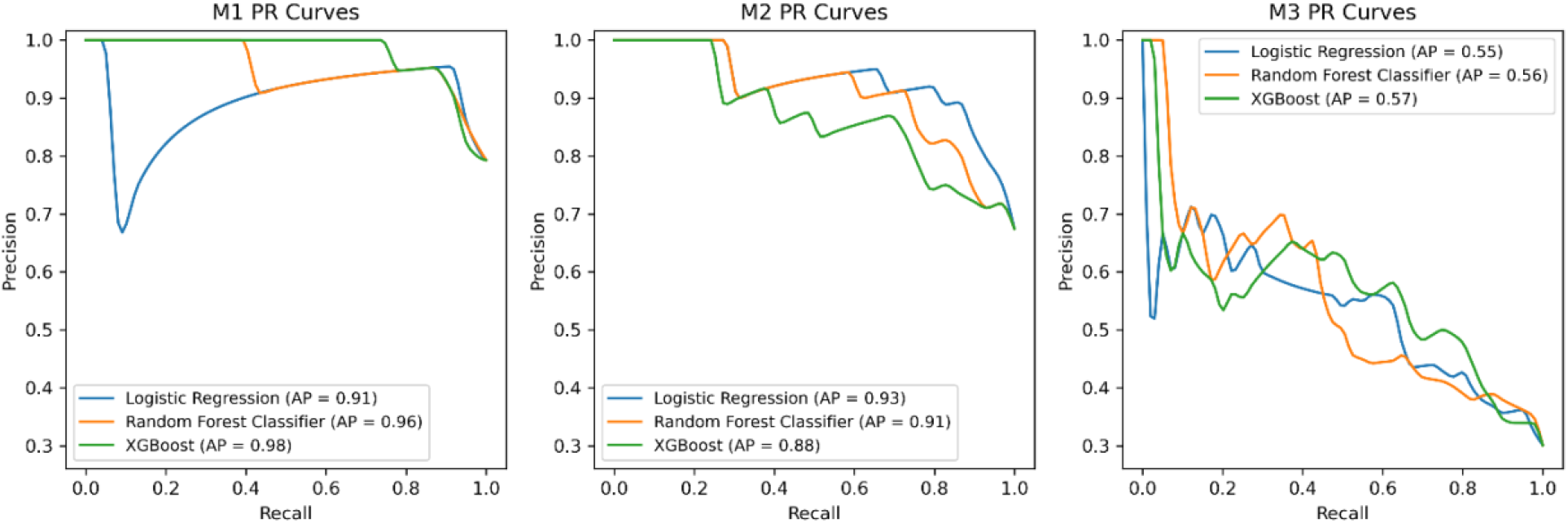
Precision-recall (PR) curves, by outcome and classifier.

## DISCUSSION

Our results suggest that clinical and demographic factors can be used to predict outcomes of patients treated with OPAT. A key feature of our study was the application of machine learning methodologies to electronic health record data from a large, tertiary academic medical center. The resulting models predicted the three primary outcomes with moderate-to-high discrimination and accuracy. The type of antimicrobials being administered and the location of treatment were among the variables most consistently related to the success or failure of OPAT in our models. While these factors may not be causally related to outcomes, they could be used to identify patients who are most likely to benefit from early discharge and outpatient management of severe infections.

For the first outcome of OPAT success at 30 days, variables with the strongest association included use of an infectious disease team consultation during hospital readmission, use of vancomycin, and worsening infection. These features may all reflect the severity of the patient’s condition. Because the variables were derived from the XGBoost model, the directionality of the association of each variable with outcome cannot be determined without using a separate model. In other words, the value of the model lies in its ability to predict outcomes and not necessarily in its ability to allow users to understand the effect of its independent features.

In contrast, the best models for the other two primary outcomes—availability of follow-up data at 30 days and occurrence of an adverse event—utilized logistic regression, which provided information about directionality and effect size for each predictor, offering greater interpretability. In our M2 model, patients who were readmitted within 30 days were also more likely to be assessable at 30 days. While this relationship seems obvious, as patients in the hospital can be easily assessed, very few patients were actually in the hospital at the 30-day time point (e.g. the readmission occurred prior to 30 days). An alternate interpretation is that patients who require readmission may demonstrate more cautious behavior and willingness to attend follow up appointments or that providers may be more vigilant with these patients.

Patients treated in a home setting were more likely to attend the 30-day assessment and to have an adverse event within 30 days. Compared to other healthcare settings like nursing homes, patients treated at home were less likely to receive close supervision. However, they may be more likely to attend follow-up appointments. Patients who required follow-up imaging were less likely to attend the 30-day assessment and less likely to have an adverse outcome. Follow-up imaging may be ordered for many reasons. While it could reflect greater severity of illness, it may also be ordered to evaluate resolving infection.

A higher frequency of adverse events was also observed for patients on vancomycin, daptomycin, or piperacillin-tazobactam. These antimicrobials are typically used for more severe infections, which likely accounts for the association rather than any pharmacologic characteristic of these drugs that makes them less amenable to outpatient (as opposed to inpatient) therapy.

Prior retrospective studies have identified a low burden of comorbid illnesses, adequacy of post-discharge follow up, and absence of intravascular infection as predictors of successful completion of OPAT therapy.^9-13^ A distinctive feature of the current study was the relatively “unbiased” selection of potential predictors, enabled by the use of machine learning approaches. While the predictors identified by our models may reflect similar underlying risk factors suggested by prior studies, e.g. severity of the patient’s illness, the use of these predictors may lead to better model performance and accuracy than pre-specified risk factors. Also, in contrast to our study, many prior studies of OPAT outcomes were conducted outside the United States, in countries with single payer systems.

Our study has several limitations that deserve discussion. Because OPAT treatment is relatively new, only a moderate number of patients have been treated at any hospital, and many patients had incomplete data. The development of more structured documentation should result in improvements in OPAT data collection over time. Smaller datasets can have significant variation between train-test splits, resulting in a higher risk of overfitting. Additionally, to fill missing values, we made several assumptions which may have reduced the effect size associated with certain features.

Our models demonstrate that clinical and demographic factors can predict outcomes for patients receiving OPAT treatment. This information should help to identify which patients might be poor candidates for OPAT or require closer observation with this approach. Conversely, identifying good candidates for OPAT offers the opportunity to shorten costly or inconvenient inpatient hospital stays.

## Data Availability

All data produced in the present study are available upon reasonable request to the authors

## References

1. Kesharwani D, Bista A, Singh H, Unnithan A, Das G, Bristoll S, Lewis N, Alnoori N. Outpatient Parenteral Antimicrobial Therapy Practice in United Kingdom: A Single-center Experience. Oman Med J. 2022;37:e442.

2. Fernandez-Polo A, Ramon-Cortes S, Plaja-Dorca J, Bartolome-Comas R, Vidal-Valdivia L, Soler-Palacin P, Grupo T-P. Impact of an outpatient parenteral antimicrobial treatment (OPAT) as part of a paediatric-specific PROA program. Enferm Infecc Microbiol Clin (Engl Ed). 2023;41:230–234.

3. Bugeja SJ, Stewart D, Vosper H. Clinical benefits and costs of an outpatient parenteral antimicrobial therapy service. Res Social Adm Pharm. 2021;17:1758–1763.

4. Thijs L, Quintens C, Vander Elst L, De Munter P, Depypere M, Metsemakers WJ, Vles G, Liesenborghs A, Neefs J, Peetermans WE, Spriet I. Clinical Efficacy and Safety of Vancomycin Continuous Infusion in Patients Treated at Home in an Outpatient Parenteral Antimicrobial Therapy Program. Antibiotics (Basel). 2022;11.

5. Durojaiye OC, Bell H, Andrews D, Ntziora F, Cartwright K. Clinical efficacy, cost analysis and patient acceptability of outpatient parenteral antibiotic therapy (OPAT): a decade of Sheffield (UK) OPAT service. Int J Antimicrob Agents. 2018;51:26–32.

6. Mahoney MV, Childs-Kean LM, Khan P, Rivera CG, Stevens RW, Ryan KL. Recent Updates in Antimicrobial Stewardship in Outpatient Parenteral Antimicrobial Therapy. Curr Infect Dis Rep. 2021;23:24.

7. Barr DA, Semple L, Seaton RA. Outpatient parenteral antimicrobial therapy (OPAT) in a teaching hospital-based practice: a retrospective cohort study describing experience and evolution over 10 years. Int J Antimicrob Agents. 2012;39:407–13.

8. Harris PA, Taylor R, Thielke R, Payne J, Gonzalez N, Conde JG. Research electronic data capture (REDCap)--a metadata-driven methodology and workflow process for providing translational research informatics support. J Biomed Inform. 2009;42:377–81.

9. Durojaiye OC, Morgan R, Chelaghma N, Kritsotakis EI. Clinical predictors of outcome in patients with infective endocarditis receiving outpatient parenteral antibiotic therapy (OPAT). J Infect. 2021;83:644–649.

10. Salles TCG, Cerrato SG, Santana TF, Medeiros EA. Factors associated with successful completion of outpatient parenteral antibiotic therapy in an area with a high prevalence of multidrug-resistant bacteria: 30-day hospital admission and mortality rates. PLoS One. 2020;15:e0241595.

11. Erba A, Beuret M, Daly ML, Khanna N, Osthoff M. OPAT in Switzerland: single-center experience of a model to treat complicated infections. Infection. 2020;48:231–240.

12. Means L, Bleasdale S, Sikka M, Gross AE. Predictors of Hospital Readmission in Patients Receiving Outpatient Parenteral Antimicrobial Therapy. Pharmacotherapy. 2016;36:934–9.

13. Cervera C, del Rio A, Garcia L, Sala M, Almela M, Moreno A, Falces C, Mestres CA, Marco F, Robau M, Gatell JM, Miro JM, Hospital Clinic Endocarditis Study G. Efficacy and safety of outpatient parenteral antibiotic therapy for infective endocarditis: a ten-year prospective study. Enferm Infecc Microbiol Clin. 2011;29:587–92.

